# Scalable In-hospital Decontamination of N95 Filtering Facepiece Respirator with a Peracetic Acid Room Disinfection System

**DOI:** 10.1101/2020.04.24.20073973

**Authors:** Amrita R. John, Shine Raju, Jennifer L. Cadnum, Kipum Lee, Phillip McClellan, Ozan Akkus, Sharon K. Miller, Wayne Jennings, Joy A. Buehler, Daniel F. Li, Sarah N. Redmond, Melissa Braskie, Claudia K. Hoyen, Curtis J. Donskey

## Abstract

**Background:** Critical shortages of personal protective equipment (PPE) especially N95 respirators during the SARS-CoV-2 pandemic continues to be a source of great concern among health care workers (HCWs). Novel methods of N95 filtering facepiece respirator (FFR) decontamination that can be scaled-up for in-hospital use can help address this concern and keep HCWs safe.

**Methods:** A multidisciplinary pragmatic study was conducted to evaluate the use of an ultrasonic room high-level disinfection system (HLDS) that generates aerosolized peracetic acid (PAA) and hydrogen peroxide for decontamination of large numbers of N95 respirators. A cycle duration that consistently achieved disinfection of N95 respirators (defined as ≥6 log_10_ reductions in bacteriophage MS2 and *Geobacillus stearothermophilus* spores inoculated onto respirators) was identified. The treated masks were then assessed for changes to their hydrophobicity, material structure, strap elasticity, and filtration efficiency (FE). Assessment of PAA and hydrogen peroxide off-gassing from a treated mask was also performed.

**Results:** The PAA room disinfection system was effective for the decontamination of N95 respirators in a 2447 cubic feet room with deploy and dwell times of 16 and 32 minutes respectively, and a total cycle time of 1 hour and 16 minutes. After 5 treatment cycles, no adverse effects were detected on filtration efficiency, structural integrity, or strap elasticity. There was no detectable off-gassing of PAA and hydrogen peroxide from the treated masks 20 and 60 minutes after the disinfection cycle respectively.

**Conclusion:** The PAA room disinfection system provides a rapidly scalable solution for in-hospital decontamination of large numbers of N95 respirators to meet the needs of HCWs during the SARS-CoV-2 pandemic.

## INTRODUCTION

The Severe Acute Respiratory Syndrome Coronavirus 2 (SARS-CoV-2) pandemic has revealed several inadequacies within healthcare. One of them has been the critical shortage of personal protective equipment (PPE) for healthcare workers (HCWs) on the frontlines of the pandemic response.^1,2^ Singleuse disposable PPE such as N95 Filtering Facepiece Respirators (FFR) and surgical facemasks are being worn for extended periods or are reused until they become soiled or visibly damaged. Shortages of PPE have been detrimental to the morale of HCWs and places them at risk for infection, death and disability.^3-5^ Given these extraordinary challenges, it has become vital to devise novel alternative methods to reuse PPE safely and effectively.

Among all PPE, the critical shortage of N95 FFRs has been most pronounced.^6,7^ At the onset of the outbreak, the Centers for Disease Control and Prevention (CDC) had recommended N95 FFRs to be used for all interactions with confirmed or suspected COVID-19 patients. It has subsequently modified its guidance regarding PPE required while caring for patients with SARS-CoV-2.^8^ At present, both the CDC and the World Health Organization (WHO) recommend the use of N95 FFRs for all aerosol-generating procedures (AGP) performed on confirmed COVID-19 patients and persons under investigation (PUI).^9,10^ Given the shortage of N95 respirators, the CDC has provided updated guidance for extended use and limited reuse of these respirators by HCWs.^11^ Several strategies have been proposed for conserving the supply of PPE including repurposing other devices to be used as FFRs, creating FFRs at home and decontamination of N95s using methods such as ultraviolet-C germicidal irradiation, dry heat, moist heat, and vaporized hydrogen peroxide.^12-15^ Vaporized hydrogen peroxide (VHP) has provided the most promising results and was recently given provisional US Food and Drug Administration (FDA) Emergency Use Authorization (EUA) for decontamination of used N95 respirators.^16^ Although the availability of VHP decontamination is an important development, the process is labor and time-intensive due to a long treatment cycle, may be limited in its access and requires the shipment of used N95 respirators to and from a central processing center.^17^ The FDA has since also granted EUA for other hydrogen peroxide based sterilization devices which are currently in use in several hospitals and allows for in-hospital disinfection of the used N95 FFRs, however these devices are limited by the number of N95 FFRs that can be processed at a given time.

There remains an urgent need for an effective N95 respirator disinfection process that will allow on-site reprocessing with rapid turnaround times, ease of use with existing personnel and expertise, and flexibility and scalability to process large quantities of respirators. Ideally, the process should allow for the disinfection of other PPE, such as gowns, surgical face masks, powered air-purifying respirator (PAPR) hoods, and face shields as the needs arise. It has been reported that aerosolized peracetic acid (PAA) and hydrogen peroxide was effective for disinfection of N95 respirators.^18,19^ Here, we expanded on these promising findings by evaluating the use of the same technology on a much larger scale for whole-room disinfection of a large number of respirators and evaluated the impact of the treatment on mask performance.

## METHODS

A multi-institutional study was conducted by researchers at University Hospitals Cleveland Medical Center (UHCMC), Case Western Reserve University (CWRU), National Aeronautical and Space Administration (NASA) Glenn Research Center and the Cleveland Veterans Affairs Medical Center (VAMC) to evaluate the use of an ultrasonic room disinfection system that generates aerosolized PAA and hydrogen peroxide for disinfection of large numbers of N95 respirators.

### Protection of Human Research Participants

The proposed PAA disinfection experiments were approved by an internal safety review at University Hospitals Cleveland Medical Center (UHCMC). The microbiologic analyses were approved by the Biosafety Committee at the VAMC. Institutional Review Board approval was not obtained as human subjects were not enrolled in the study.

### Development and Optimization of the PPE Decontamination Room

The PAA room high-level disinfection system (HLDS) (AP-4™, Altapure, Mequon, WI) was placed in the center of a 16.3 x 16 x 9.5 feet room (2447 cu. ft) (**Figure 1A-E**). The device uses ultrasonic vibrations to generate a dense cloud of submicron droplets of PAA, each containing peracetic acid (0.18%), hydrogen peroxide (0.88%), water (98.58%); and the remainder being inert ingredients.^20^ The aerosol eventually decomposes into non-toxic end products: water vapor, acetic acid (vinegar) and oxygen. The decontamination cycles consisted of 4 phases: Deploy (deployment of the PAA submicron aerosols into the room), Dwell (aerosols left to stand in the room), Scrub (clearance of the aerosol by dehumidification and by drawing it through activated charcoal filters) and Vent (fresh air is circulated through the room by opening the manual vents enabling clearance of residual vapors and drying of the masks). The ventilation in the test room was modified to allow the influx and circulation of fresh air at the end of the scrub cycle. An extra air scrubber (HJ-200™, Altapure, Mequon, WI) was deployed to keep vent times to a minimum by accelerating clearance of residual PAA. The deploy and dwell times are directly responsible for microbial reduction, whereas the scrub and the vent cycles impact clearance of residual PAA vapors to recommended safe levels. There are no specific OSHA standards for PAA.^21^ The American Conference of Governmental Hygienists (ACGIH) has set a Threshold Limit Value (TLV) of 0.4 ppm, as a 15-minute Short Term Exposure Limit (STEL).^21-23^ The U.S. Environmental Protection Agency (EPA) recommended Acute Exposure Guidelines (AEGL-1) limit is 0.17 ppm (0.52 mg/m^2^).^24^

**Figure 1.**
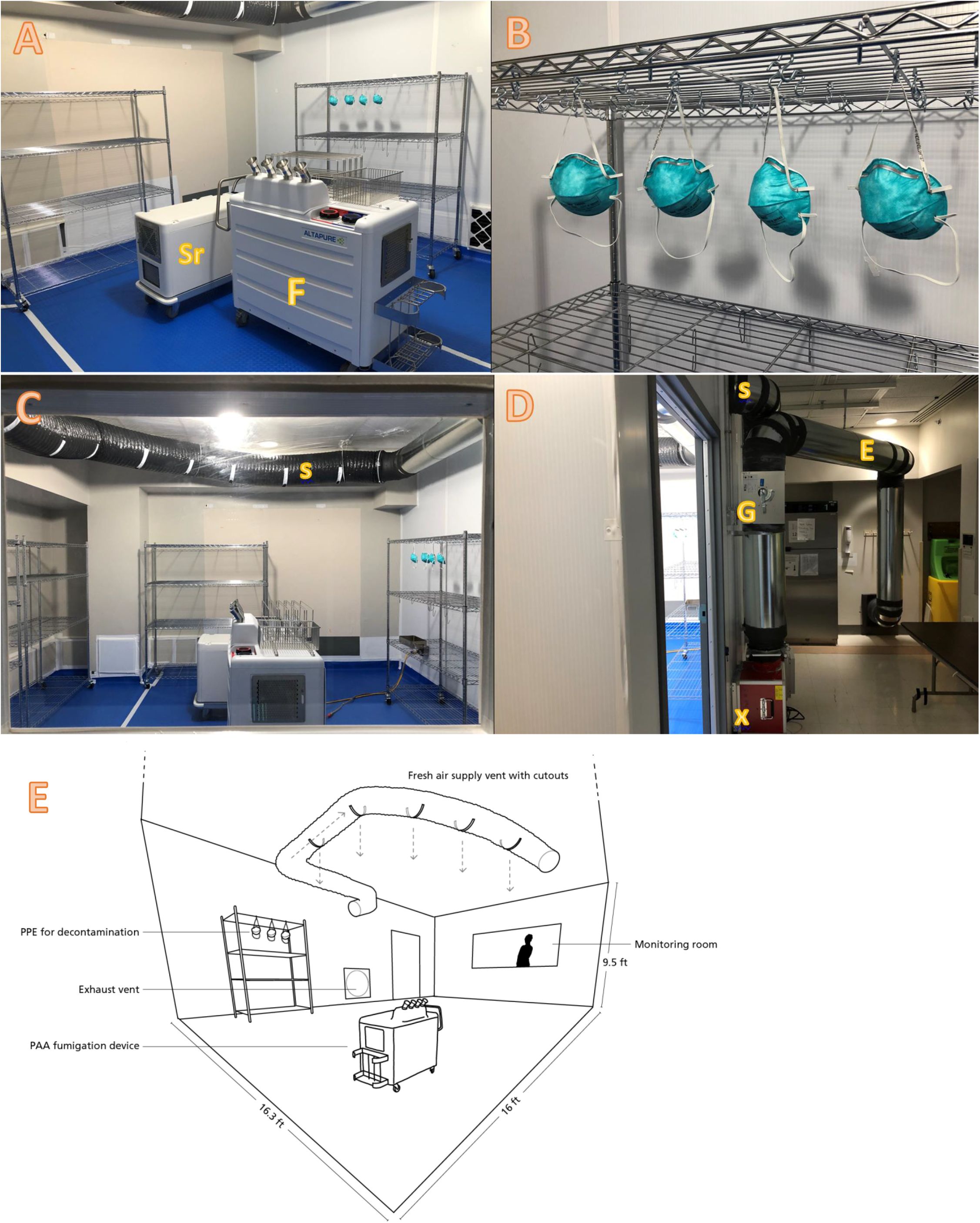
shows A) Placement of the aerosolization device (F) and extra scrubber (Sr) in the middle of the test room B) N95 masks suspended on ‘S’ shaped hooks C) Test room layout with ventilation on the ceiling providing fresh air (supply) into the room during the vent cycle D) the ventilation set up with supply (S) and exhaust (E); note two 600 cfm blower fans (X) in a push-pull configuration with manually operated gasketed dampers (G). E) Schematic diagram of the room dimensions and ventilation system.

Before the start of the disinfection cycle, the deploy, dwell, scrub and vent phase times were manually configured using the application programming interface (API) on the accompanying digital tablet (**Figure 2**), which in turn, remotely connected to the HLDS using Bluetooth® technology. The deploy and the dwell times were adjusted to provide effective disinfection of the masks with the least possible amount of exposure to the PAA. The PAA concentrations in the room were measured in real-time by a PAA sensor (Safecide™, ChemDAQ, Pittsburgh, PA). At the end of the vent cycle, PAA concentrations were at 0.12 ppm, below the AEGL-1 limit of 0.17 ppm **(Figure 3)**. The 15-minute time-weighted average (TWA) of PAA concentration after the decontamination cycle was 0.08 ppm (15-min STEL for PAA is 0.4 ppm).

**Figure 2.**
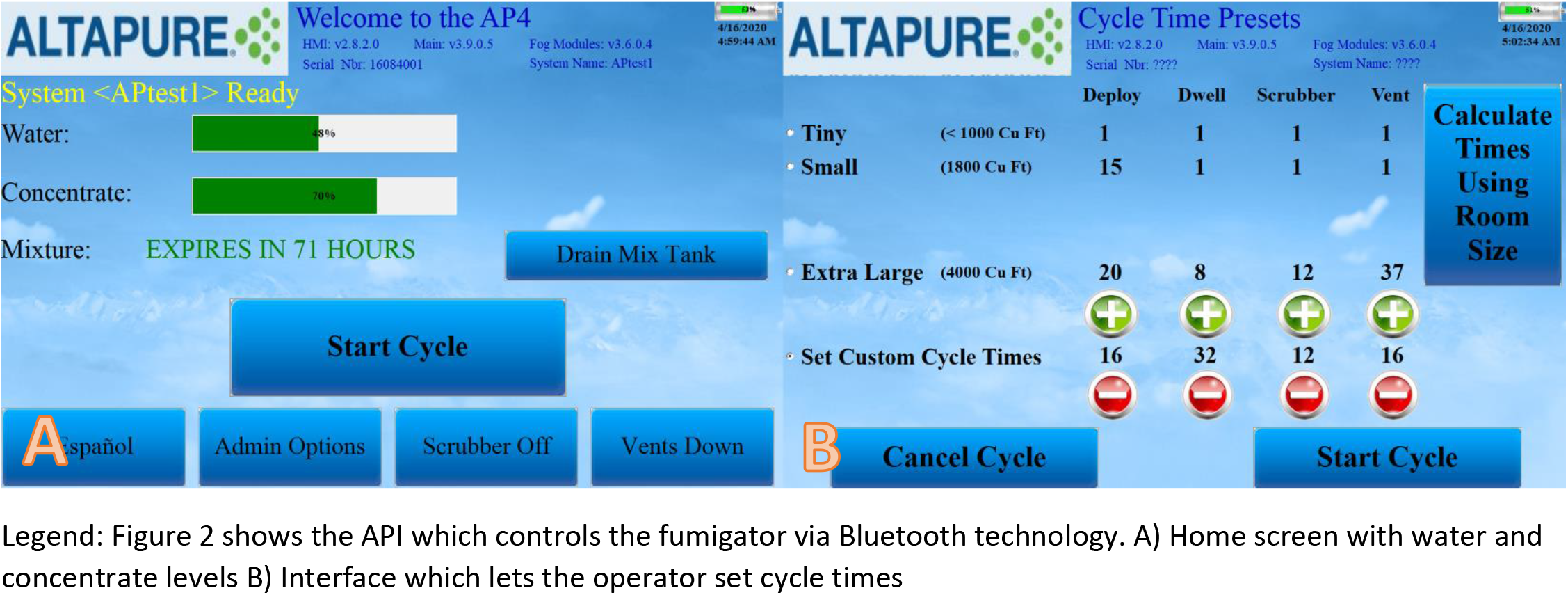
shows the API which controls the fumigator via Bluetooth technology. A) Home screen with water and concentrate levels B) Interface which lets the operator set cycle times

**Figure 3.**
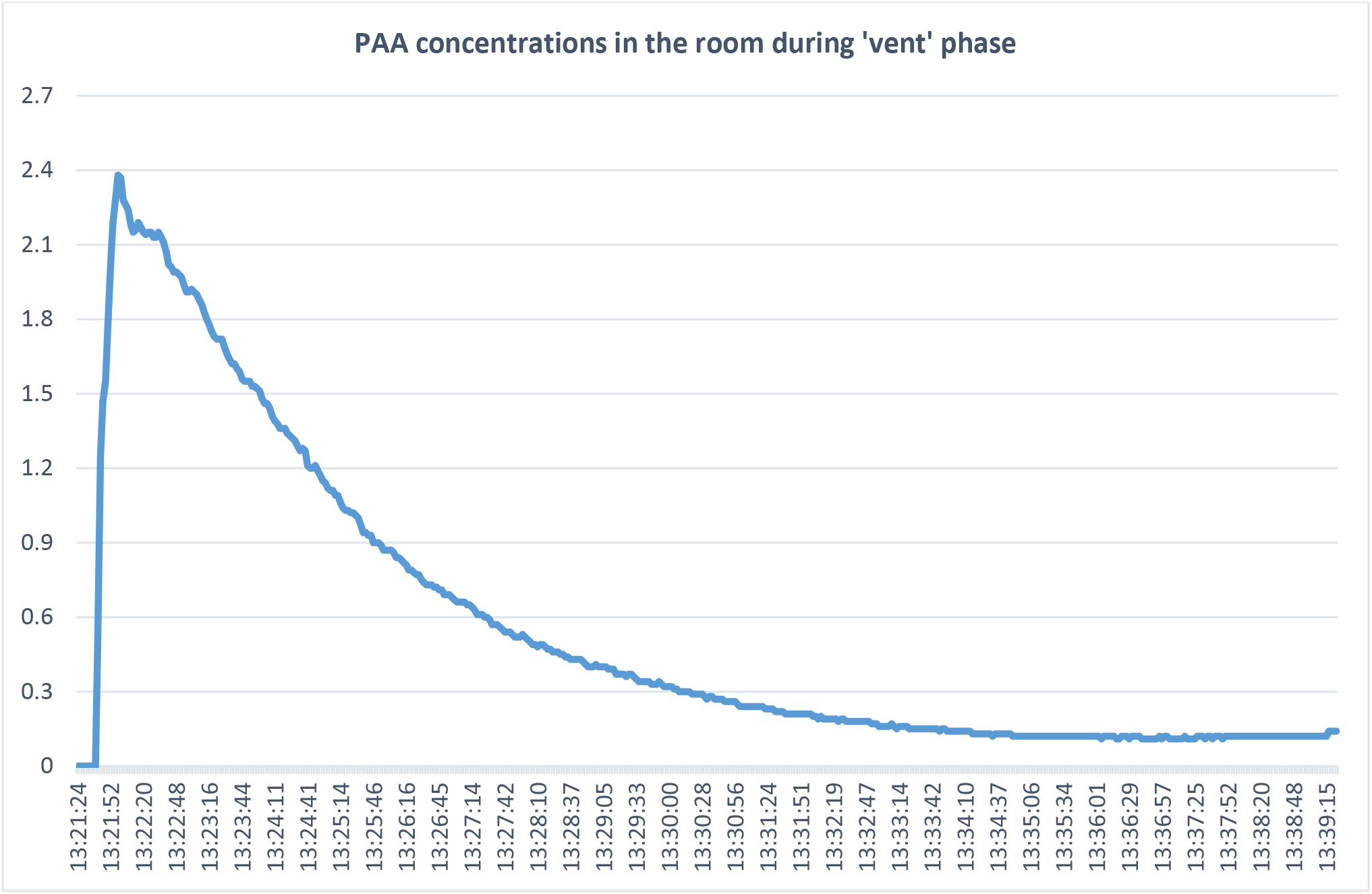
shows the PAA concentrations in the test room at the beginning of the vent phase when the blower fans are turned on (active ventilation). PAA concentrations in the room drop to 0.12 ppm after 16 minutes. (The AEGL-1 limit for PAA over a 10 min or 8 hour period is 0.17 ppm)

Three test cycles were evaluated to identify a cycle time achieving consistent disinfection of bacteriophage MS2 and *G. stearothermophilus* spores inoculated onto N95 respirators. The deploy time on these test cycles ranged from 12 minutes (default settings for a room of 2447 cu. ft) to 19 minutes. The shortest cycle tested was 44 minutes (deploy: 12 minutes, dwell: 8 minutes, scrub: 8 minutes and vent: 16 min). Based on the results, the cycle times were incrementally adjusted to get an optimal cycle time. The optimal cycle was identified as being the shortest cycle at which disinfection was consistently achieved (deploy: 16 minutes, dwell: 32 minutes, scrub: 12 minutes and vent: 16 minutes). This cycle was then repeated up to 5 times with sterile masks that were then sent for analysis of structural integrity, instantaneous filtration efficiency and load testing analysis.

### Efficacy of the decontamination process for treatment of contaminated N95 respirators

The model 1860 N95 (3M, Minneapolis, USA) respirator was studied because it was the respirator used at the study hospital. Twenty new respirators were tested from 2 different lots available from the hospital inventory. Two N95 respirators were used for each decontamination test cycle. The test and control respirators were inoculated with ~10^6^ colony-forming units (CFU) of *G. stearothermophilus* spores and ~10^6^ plaque-forming units (PFU) of bacteriophage MS2 on the outer and inner surface of the respirator using previously described methods.^18,25,26^ The test organisms suspended in 8% simulated mucus,^27^ and 10 μL aliquots were pipetted onto the respirator surface and spread with a sterile loop to cover an area of 1 cm^2^ and allowed to air dry. The test N95 respirators were suspended using metallic ‘S’ shaped hooks from metal wire shelving carts at a height of 6 feet (height of the tallest shelf) and exposed to PAA submicron droplets. The control masks were left untreated at room temperature, maintained at 68°F.

After the disinfection treatments, the inoculated sections of the N95 respirators were cut out, vortexed for 1 minute in 1 mL of phosphate-buffered saline with 0.02% Tween and serial dilutions were plated on selective media to quantify viable organisms. Broth enrichment cultures were used to assess for recovery of low levels of *G. stearothermophilus* spores. All tests were performed in triplicate. Log_10_ CFU or PFU reductions were calculated by comparing recovery from treated versus untreated control respirators.

### Evaluation of Contact Angle on the Surface of Treated N95 Respirators

The contact angle on the surface of untreated and treated N95 respirators was measured with a Kernco Instruments contact angle meter at the NASA Glenn Research Center. A micropipette was used to place a small droplet of deionized water on the surface (outer green layer) of a ~1/2”x1” section cut from each mask with scissors. Contact angle (θ) (**Figure 4**) for each of 3 drops was measured using the Goniometer scale on the instrument for each sample. The range of angles measured was documented (**Table 1**).

**Figure 4.**
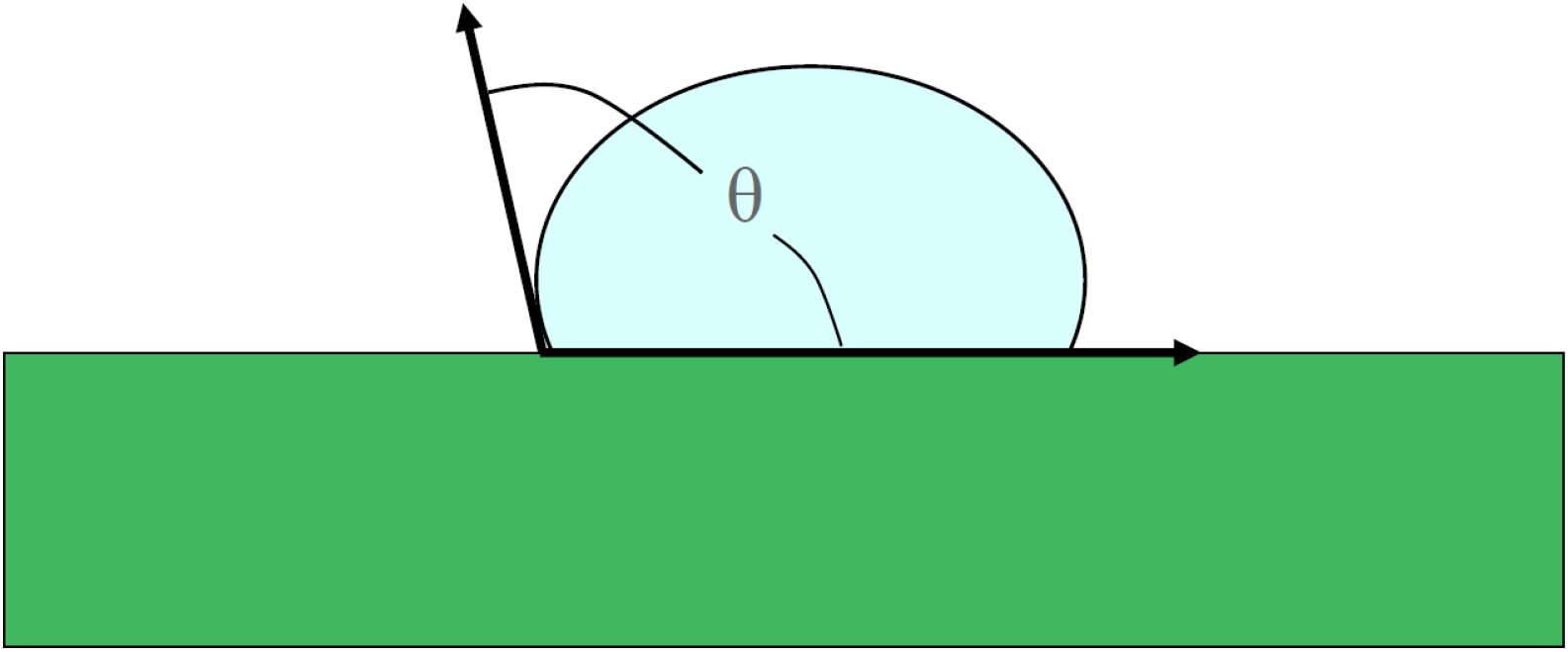
demonstrates measurement of the contact angle during hydrophobicity testing.

**Table 1.** shows the contact angle with repeated cycles of PAA disinfection. The cycle length was a dwell of 12 minutes and a deploy of 8 minutes. ‘N’x indicates number of times the N95 FFR was treated with this cycle.

| No of cycles | Contact angle (Degrees) |
| --- | --- |
| Control Mask (0 runs) | 97-99 |
| 1x | 97-99 |
| 2x | 97-99 |
| 3x | 97-99 |
| 4x | 97-99 |

### Evaluation of N95 Respirator Structural Integrity by Scanning Electron Microscopy

The outer (green) fabric of the mask was examined using scanning electron microscopy (SEM) at the NASA Glenn Research Center. Samples of approximately 1”x1” cut from each mask with scissors were coated with a 10 nm layer of platinum to reduce charging in the electron beam and then mounted to a 4” pin-mount platen with conductive carbon tape for SEM viewing. A Tescan MAIA-3 Scanning Electron Microscope was used to view the fibers in each mask sample. The test parameters were: accelerating voltage, 1 kV; working distance, 15 mm; beam intensity, set at 8 resulting in an absorbed current of about 180pA; and spot size, ~28 nm.

### Effect of treatment on elasticity of the respirator straps

Three samples were cut from elastic straps of two masks at each sterilization cycle (0, 3 and 5 cycles), resulting in a sample size of 6 per group. Samples were 3 cm in length and were mounted to the grips of a tensile testing machine. The gage length of specimens was set at 1 cm length. Samples were loaded under displacement control (800L, Testresources, Minnetonka, MN) at a rate of 1 mm/sec. The testing profile included two cycles of load relaxation such that in each cycle the samples were loaded to 300% strain (i.e. to net length of 30 mm following 20 mm deformation), with samples held at constant deformation for 5 minutes. There was a 5-minute period between the end of the first cycle and the beginning of the second cycle. The load attained at 300% strain was obtained from the curves for each cycle as ‘Load 1’ and ‘Load 2’ values. Load-relaxation was calculated for each cycle as the difference of the loads at the beginning and end of the load period as ‘Relaxation 1’ and ‘Relaxation 2’. As such, relaxation represents the capacity of straps to retain a load over time. The elasticity of samples was calculated as the secant stiffness from the loading portion of cycle 1 and defined as ‘Stiffness 1’. Secant stiffness was the slope of the line connecting the origin with the point that is defined as Load 1. Owing to the limited sample size, a non-parametric Kruskal-Wallis test was used for one-way analysis of variance (ANOVA) at a significance level of *P*<0.05.

### Filtration efficiency of the N95 masks following exposure to PAA vapor

Evaluation of filtration efficiency was performed at ICS Laboratories, Inc. (Brunswick, OH). N95 respirators subjected to multiple runs of the optimal cycle were subjected to testing for instantaneous filtration efficacy in accordance with NIOSH standard TEB-APR-STP-0059.^28^ The masks were conditioned at 85% +/-5% relative humidity and 38°C +/-2°C for 25 hours before the filter efficiency test. Each mask was then assembled into a fixture and subjected to instantaneous aerosol loading. The sodium chloride aerosol was neutralized to a Boltzmann equilibrium state at 25 +/-5°C and relative humidity of 30 +/-10%. Particle size distribution was verified to be a count median diameter of 0.075 +/-0.020 microns, with a geometric standard deviation not exceeding 1.86. The loading was performed by depositing sodium chloride aerosol at an airflow rate of 85 L/min The flow rate, initial resistance, and initial penetration data were recorded.

### Measurement of peracetic acid and hydrogen peroxide off-gassing after disinfection

Following the optimal disinfection cycle, an N95 FFR was taken out of the decontamination room and allowed to air dry in an adjacent room with a fan blowing at 500 cu. ft per minute (cfm). The N95 FFRs were tested for off-gassing after the optimal disinfection cycle and at 20-minute intervals. Testing was concluded once 2 consecutive tests showed no off-gassing for PAA and hydrogen peroxide.

The N95 FFR was placed in a sealed polyvinyl chloride cylinder (0.35 cu. ft) with airflow at 1.5L/min entering through one end and a peracetic acid or hydrogen peroxide sensor (Safecide™, ChemDAQ, Pittsburgh, PA) connected to the other end. **(Figure 5)** A 15-minute TWA for peracetic acid or hydrogen peroxide exposure was measured.

**Figure 5.**
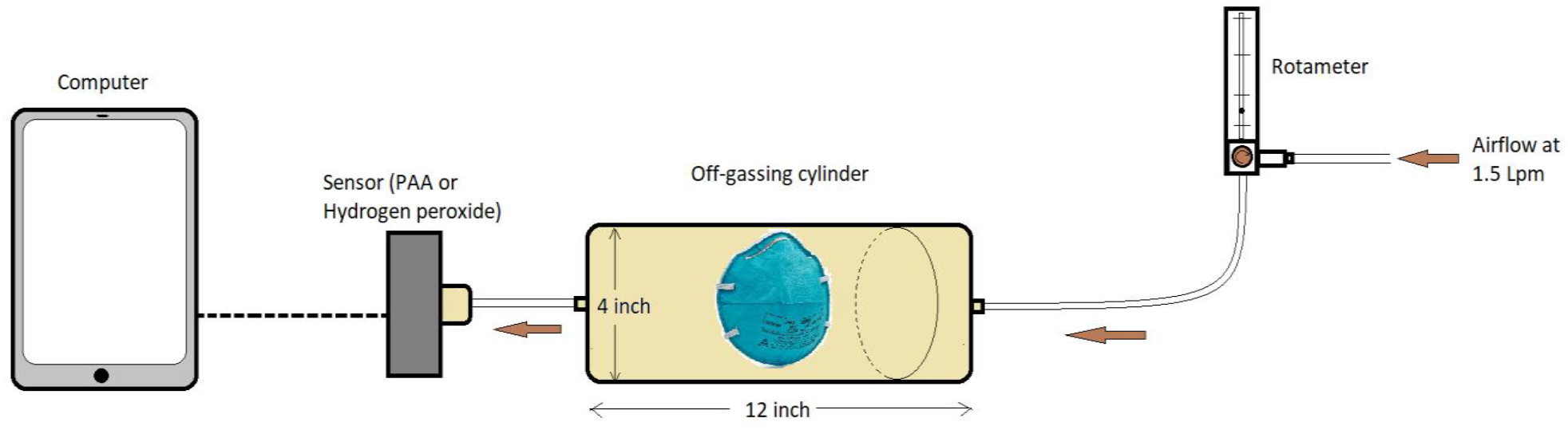
shows the off-gassing set up. The treated mask was placed in a sealed cylinder and with a constant airflow of 1.5 Lpm from one end. The other end was connected to a PAA/H2O2 sensor and 15-minute TWA levels were documented.

## RESULTS

### Efficacy of PAA disinfection of N95 masks

As shown in **Figure 6**, 6 log_10_ reductions in *G. stearothermophilus* spores were achieved on inoculated 1860 N95 FFRs with all cycle durations. For bacteriophage MS2, 6 log_10_ reductions were achieved on the inoculated 1860 N95 FFRs with the 16 minute deploy and 32 minute dwell (total cycle time 76 minutes) and the 19 minute deploy and 32 minute dwell (total cycle time 87 minutes) cycles, whereas only ~4 log_10_ reductions were achieved with the 12 minute deploy and 8 minute dwell cycle. Based on these results, the optimal disinfection cycle time was determined to be a deploy and dwell of 16 and 32 minutes respectively.

**Figure 6:**
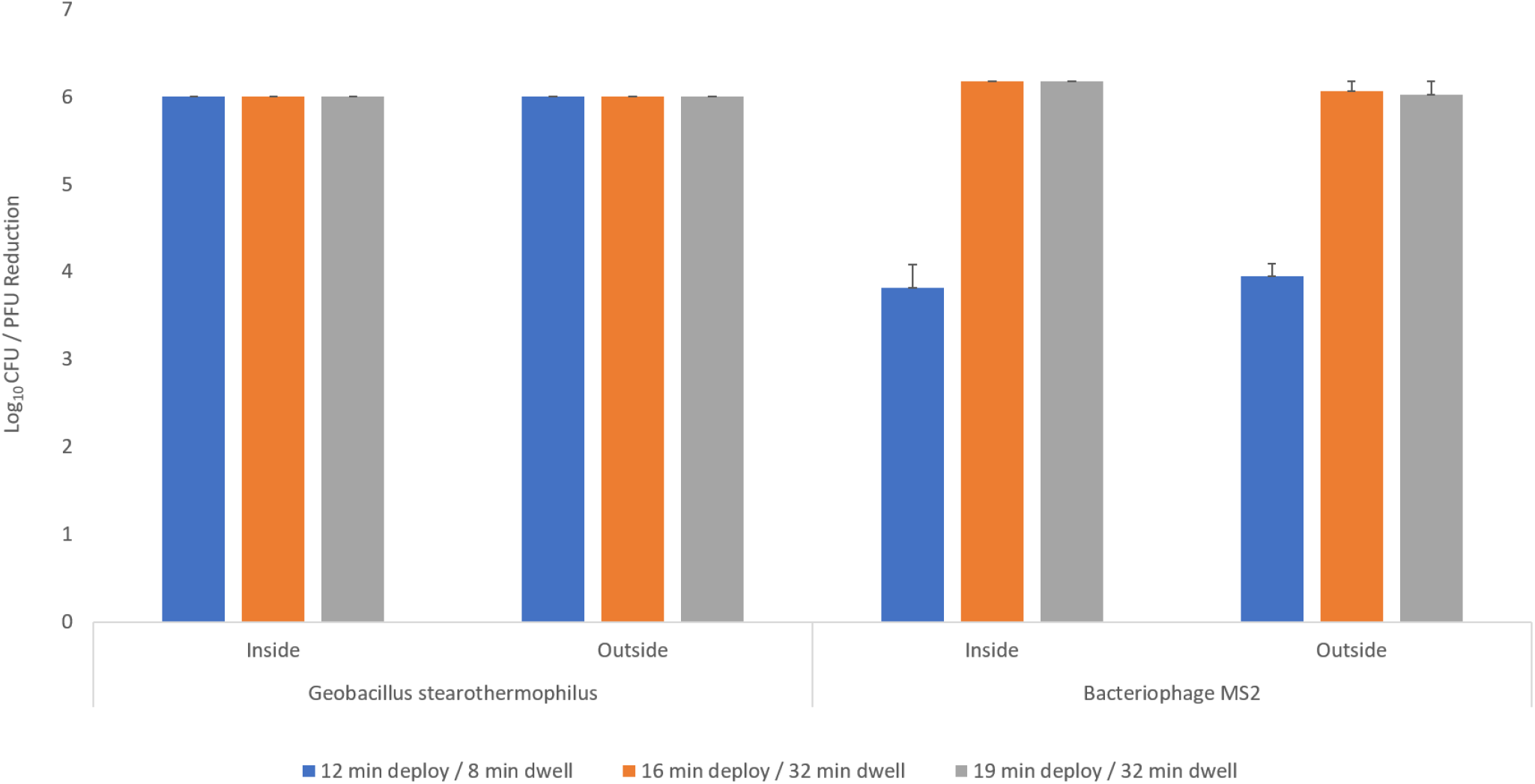
Efficacy of PAA HLDS for decontamination or disinfection of G. stearothermophilus spores and MS2.The respirator was exposed to 3 different cycles as in the figure and log_10_ reductions CFU/PFU studied. Error bars indicate standard error

### Structural integrity of the N95 masks following exposure to aerosolized PAA

SEM analysis indicated evidence of bubbles on the surface of the PAA-treated respirator outer fabric fibers which appeared to increase with the number of PAA cycles (**Figure 7A-E**). Energy Dispersive Spectroscopy dot map images of the bubble feature on PAA Cycle 4 outer mask fabric indicating that the bubbles are high in oxygen, phosphorous and nitrogen, based on the bright areas of the dot map images. The overall spectrograph shows that the surface is predominantly carbon, oxygen and phosphorous. (**Figure 8A,B**)

**Figure 7.**
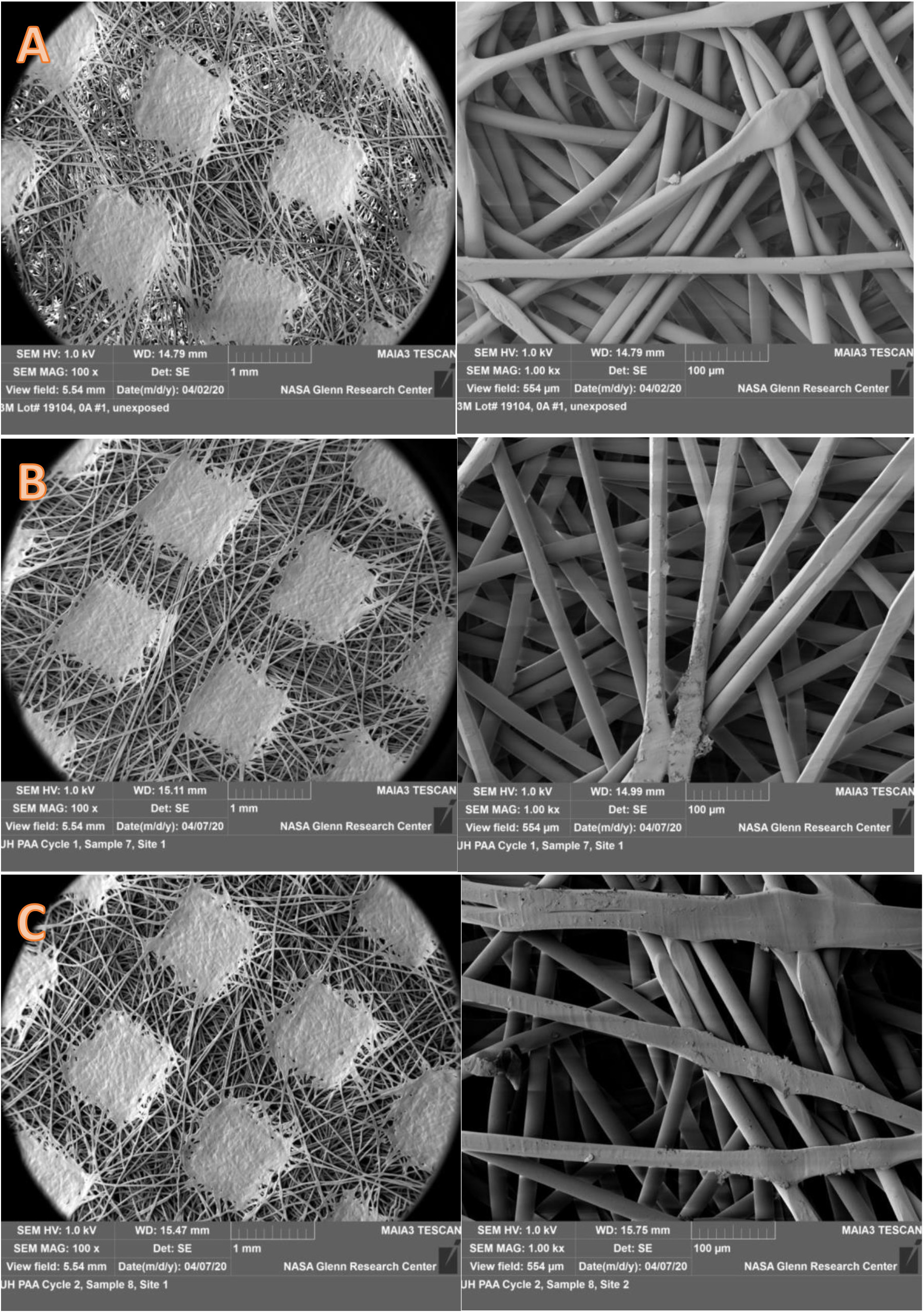
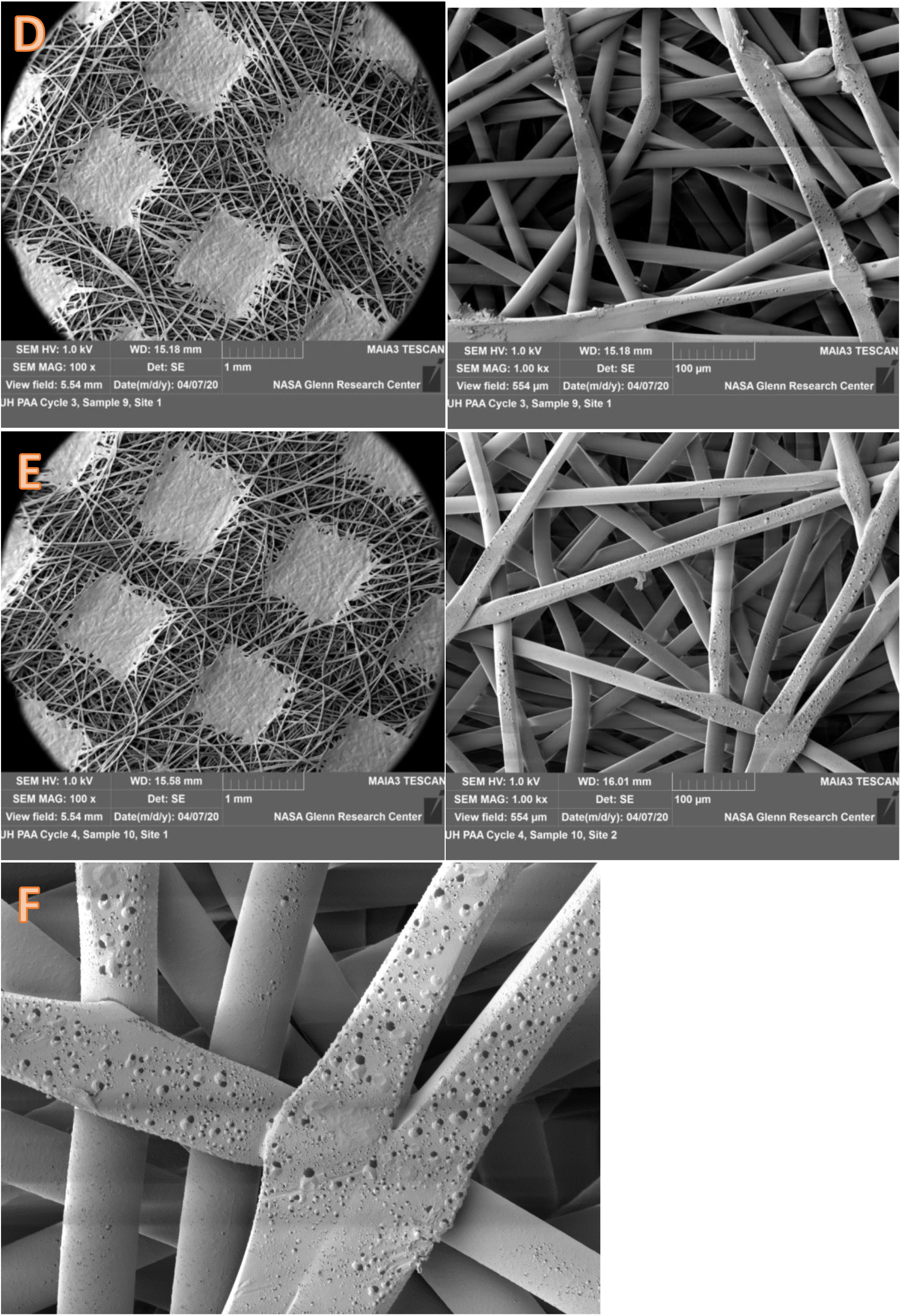
shows SEM images of the outer layer of the N95 mask under 100x (images in the left column) and 1000x magnification (images in the right column. A (control), B though E indicate multiple cycles of PAA treatment from 1-4. Note increase bubbling on the fibers after PAA exposure. Magnified image of bubbling on fibers (7F)

**Figure 8:**
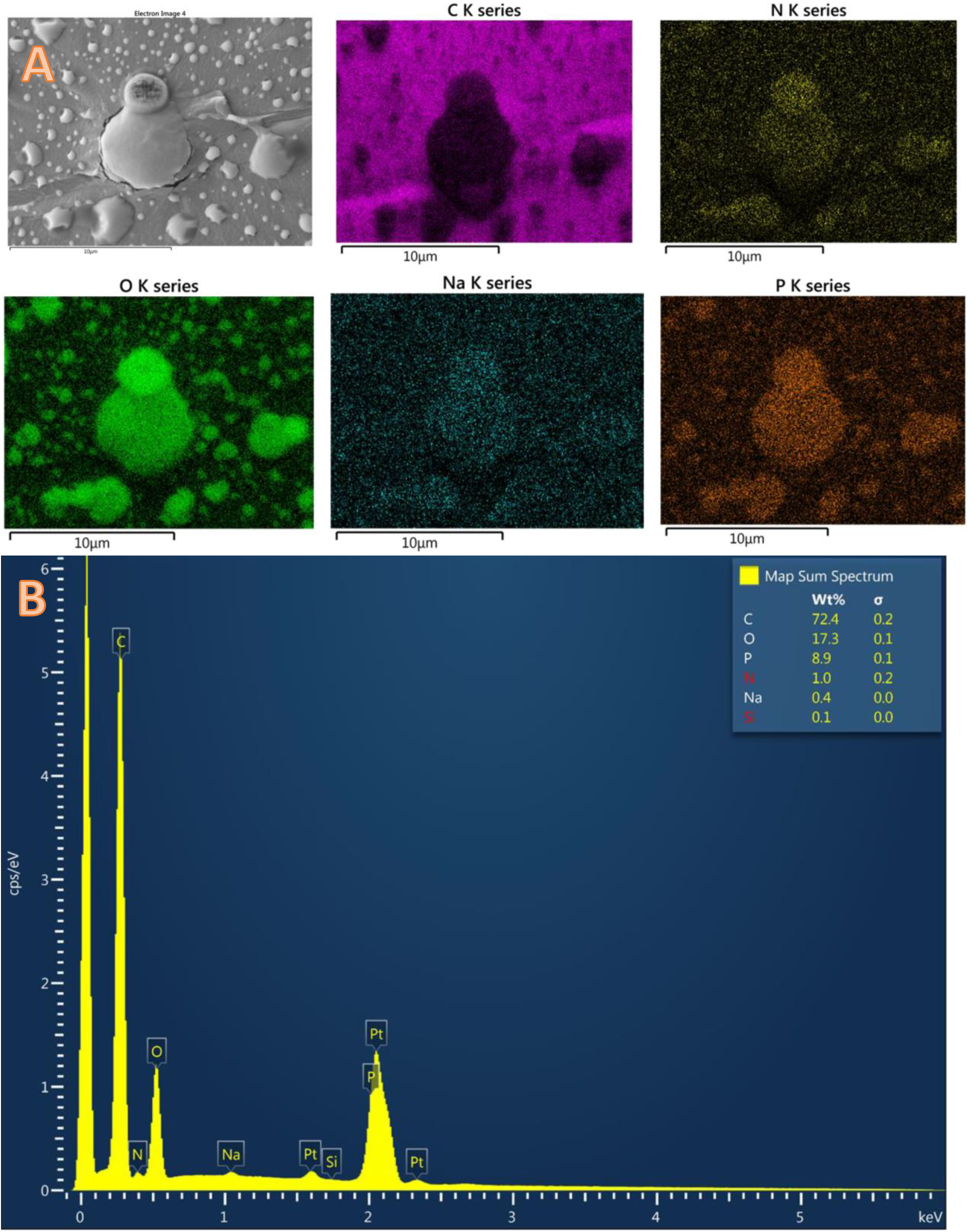
EDS showed that the bubbles are high in oxygen, phosphorous and nitrogen indicated by the bright areas of the dot map images (8A). Overall spectrograph shows that the surface is predominantly carbon, oxygen and phosphorous (8B)

### Evaluation of the contact angle on the surface of the treated N95 masks

Contact angle did not change with repeated cycles of PAA disinfection (**Table 2**), thereby concluding that the hydrophobicity of the outer layer was preserved

**Table 2.**
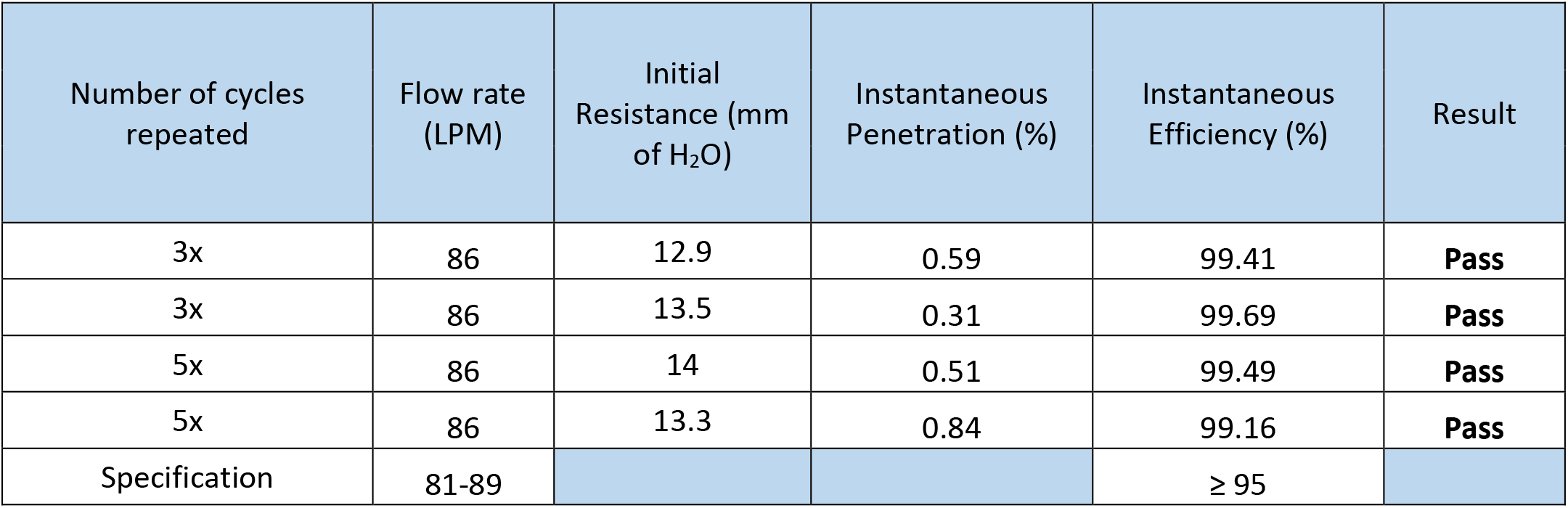
outlines the results of the instantaneous loading tests for filtration efficiency. The cycle length was a dwell of 16 minutes and a deploy of 32 minutes (optimal cycle). ‘N’x indicates number of times the N95 FFR was treated with this cycle.

| Number of cycles repeated | Flow rate (LPM) | Initial Resistance (mm of H<sub>2</sub>O) | Instantaneous Penetration (%) | Instantaneous Efficiency (%) | Result |
| --- | --- | --- | --- | --- | --- |
| 3x | 86 | 12.9 | 0.59 | 99.41 | **Pass** |
| 3x | 86 | 13.5 | 0.31 | 99.69 | **Pass** |
| 5x | 86 | 14 | 0.51 | 99.49 | **Pass** |
| 5x | 86 | 13.3 | 0.84 | 99.16 | **Pass** |
| Specification | 81-89 |  |  | ≥ 95 |  |

### Effect of treatment on elasticity of the respirator straps

None of the mechanical properties were affected significantly by the number of cycles of sterilization (P values ranged from 0.27 to 0.505). **Figure 9.A**. shows the two-cycle stress relaxation loading protocol. **Figure 9.B**. shows the load versus time profile and including the load-relaxation for each cycle calculated as the difference of the loads at the beginning and end of load period and shown as ‘Relaxation 1’ and ‘Relaxation 2’. **Figure 9.C**. shows the load versus deformation behavior over the two cycles. As such, relaxation represents the capacity of straps to retain a load over time. Elasticity of samples was calculated as the secant stiffness from the loading portion of cycle 1 and defined as ‘Stiffness 1’. Secant stiffness was the slope of the line connecting the origin with the point that is defined as Load 1 in Figure 8.C. Based on these data, elasticity, viscoelasticity and load retention capacity of the strap material were retained for up to 5 cycles (**Figure 10**).

**Figure 9:**
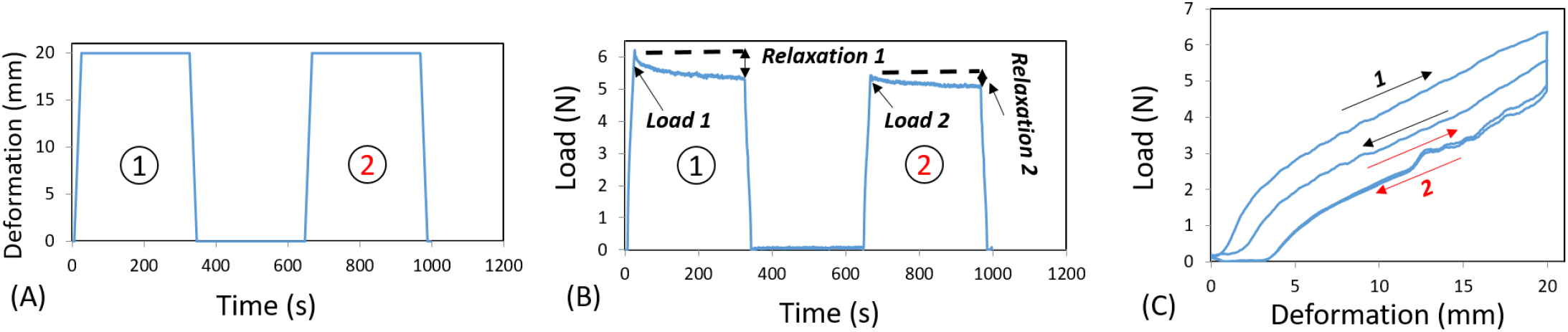
Two-cycle stress relaxation loading protocol. A) Deformation vs. time profile. B) Load vs. time profile and calculated variables. C) Load vs. deformation behavior over the two cycles.

**Figure 10:**
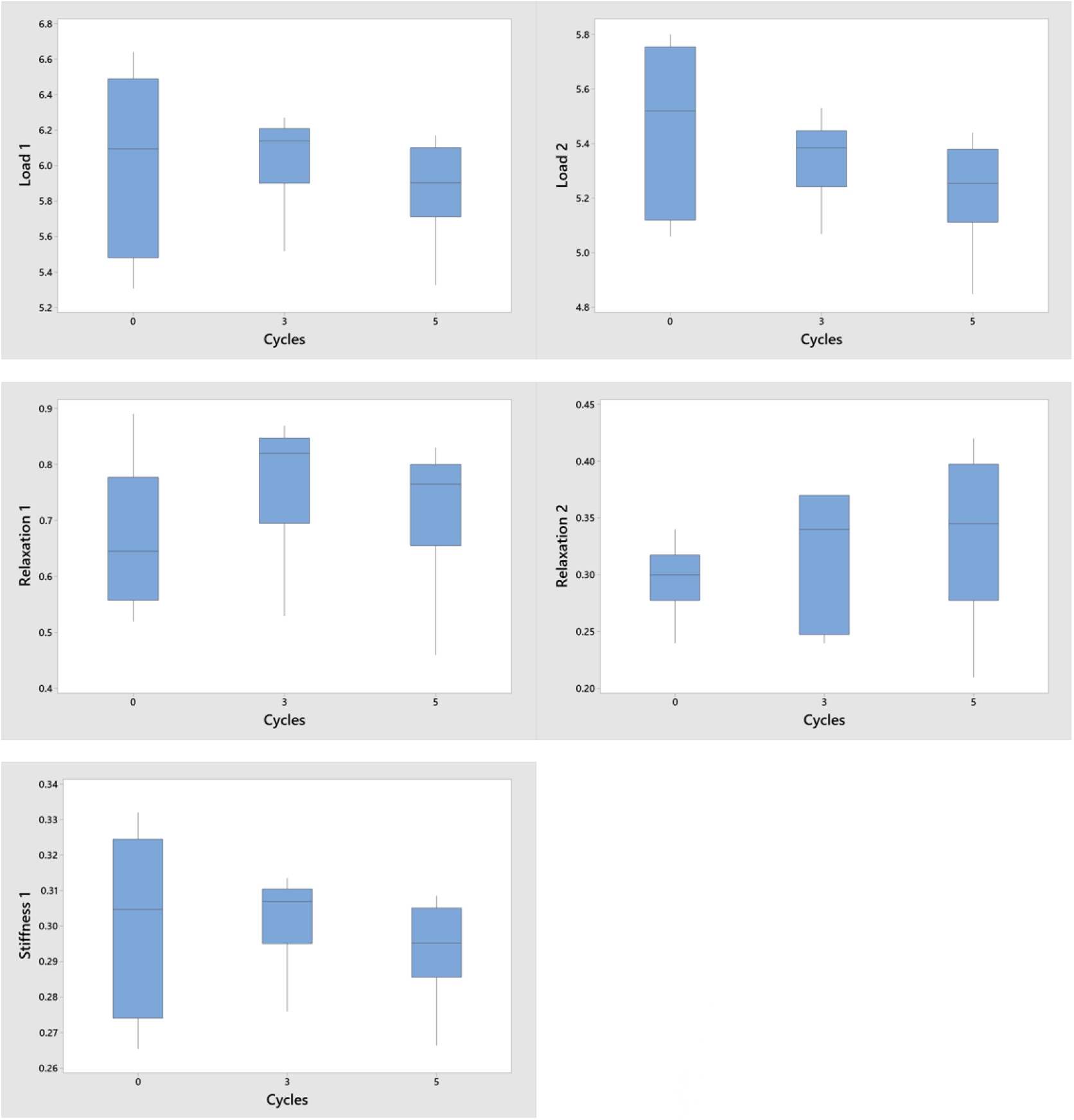
Interval plots for mechanical test variables as a function of disinfection cycles. The horizontal line is the median, the box indicates the interquartile range and the whiskers extend to the minimum and maximum values.

### Filtration efficacy of the N95 masks following exposure to PAA vapor

**Table 3** shows the results of instantaneous filtration efficiency on the masks subjected to 5 cycles of PAA treatment. There was no decline in filtration efficiency in the N95 respirators for up to 5 cycles of PAA disinfection.

**Table 3.** outlines the results of the full loading tests for filtration efficiency. The cycle length was a dwell of 16 minutes and a deploy of 32 minutes. ‘N’x indicates the number of times the N95 FFR was treated with this cycle.

| Number of cycles repeated | Flow rate (LPM) | Initial Resistance (mm of H<sub>2</sub>O) | Initial Penetration (%) | Maximum Penetration (%) | Filter Efficiency (%) | Result |
| --- | --- | --- | --- | --- | --- | --- |
| 5x | 85 | 14.9 | 0.56 | 1.35 | 98.65 | **Pass** |
| 5x | 86 | 13.8 | 0.6 | 1.37 | 98.63 | **Pass** |
| Specification | 81-89 |  |  | ≤ 5.0 | ≥ 95 |  |

### Results of PAA and Hydrogen Peroxide off-gassing after disinfection

At 20 minutes after the optimal disinfection cycle, the PAA off-gassing was measured at 0.00 ppm. At 60 minutes after the optimal disinfection cycle, the hydrogen peroxide off-gassing was measured at 0.00 ppm. **Table 4** outlines the full results of the testing.

**Table 4.** outlines the results of the hydrogen peroxide off-gassing from the N95 FFR which had undergone an ideal decontamination cycle.

| Time after cycle | Instantaneous | 15 min STEL |
| --- | --- | --- |
| 0 | 2.4 ppm | 1.76 |
| 20 min | 1.4 ppm | 1.15 |
| 40 min | 0.2 ppm | 0.18 ppm |
| 60 min | 0.0 ppm | 0.0 ppm |
| 80 min | 0.0 ppm | 0.0 ppm |

## DISCUSSION

The goal of this investigation was to address the urgent need for an effective N95 respirator decontamination process allowing onsite reprocessing with rapid turnaround times, ease of use, and scalability to process large numbers of respirators. We found that the PAA room disinfection system was easy to set up, operate and was effective for disinfection of N95 respirators with a total cycle time of 1 hour and 16 minutes. Using multiple methods, we did not detect any adverse effects on filtration efficiency, structural integrity, or strap elasticity after 5 treatment cycles. There was no detectable off-gassing of PAA and hydrogen peroxide from the treated masks after they were allowed to air dry for 60 additional minutes after the disinfection cycle.. These results suggest that the PAA room disinfection system provides a scalable solution for in-hospital decontamination of N95 respirators to meet the needs of HCWs during the SARS-CoV-2 pandemic.

Microbiological agents chosen to test for disinfection were based on guidance provided by the FDA EUA document.^29^ During the Ebola outbreak, the CDC recommended the use of disinfectants that were registered to be effective against non-enveloped viruses - as compared to enveloped viruses such as SARS-CoV-2-since they were more resistant to disinfection.^30,31^ Bacteriophage MS2 is a non-enveloped virus that has been used as a surrogate in studies looking at airborne RNA viral pathogens, as well as disinfectant studies performed by the U.S. EPA.^32-34^ *G. stearothermophilus* spores have been used in the study of PAA decontamination of surfaces.^35^

Our findings are consistent with previous studies that demonstrated the efficacy of the PAA disinfection system. Both the room HLDS and a high-level disinfection cabinet were effective in reducing pathogens, including *C. difficile* spores, on steel disk carriers by greater than 6 log_10_ CFU.^25,36^ However, an extended cycle with the disinfection cabinet was required to achieve a 6 log_10_ reduction in bacteriophage MS2 inoculated on N95 respirators.^18^ In the current study, an extended cycle (identified as the optimal disinfection cycle in our experiments) was also required to achieve a 6 log_10_ reduction in bacteriophage MS2 or *G. stearothermophilus* spores on N95 respirators. Similarly, Battelle has reported a prolonged VHP cycle time with a total time of 480 minutes for N95 decontamination.^17^

Our results demonstrate that the N95 respirators retain their structural integrity, outer surface hydrophobicity and strap elasticity for at least 5 repeated cycles of PAA treatment. However, on a microscopic level, there was evidence of visible bubbling on the non-woven polypropylene fibers of the outer layer, which increased proportionally with the duration of exposure to PAA. The significance of these bubbles is unclear at this time. It could be indicative of a trend towards loss of structural integrity with continued exposure to PAA. These changes, however, did not affect the filtration efficiency of the treated masks. Off-gassing of PAA from the treated mask was undetectable after just 20 minutes of air drying. This may be explained by the low concentrations of PAA in the aerosols, much shorter exposure times to PAA when compared to VHP and the inherently unstable nature of the compound leading to its rapid decomposition.^17,37^ The hydrogen peroxide off-gassing was found to be undetectable after 60 minutes of air drying.

The PAA room disinfection system offers several advantages over other technologies being evaluated for PPE decontamination. The technology is substantially more effective than ultraviolet-C (UVC) light for N95 decontamination.^18,38^ The aerosols allow for complete coverage of all surfaces on the masks, thus eliminating the concerns of ‘shadow areas’ with UVC germicidal irradiation.^39^ Compared to VHP, the cycle times with PAA are considerably shorter, allowing rapid turnaround times.^17,40^ This can be vital for healthcare systems to achieve decontamination of large numbers of N95 FFRs during the COVID-19 pandemic. The platform is scalable and can be replicated in real-world hospital settings. We conservatively estimate that about 2000 N95 respirators can be effectively disinfected in a room with the dimensions of the test room (2447 cu. ft), with capacity increasing proportionally to the room dimensions. An estimated 15000-20000 N95 FFRs can be decontaminated per day with this method. The disinfection room can be set up relatively easily with simple modifications to the ventilation setups in most hospital rooms. The software is easy to use and the AP-4™ can be operated by hospital personnel with minimal training. The PAA room HLDS is currently used for terminal disinfection of patient rooms in some centers across the United States and abroad. It can be readily repurposed for N95 decontamination without much added costs. While the use of VHP is not recommended with cellulose-containing materials, PAA could potentially be used with masks containing cellulose.

The PAA room disinfection system has some disadvantages. The PAA aerosols are hazardous requiring that the ventilation system is closed and the room sealed during operation. While the AP-4 HLDS is designed to disinfect rooms of varying sizes, a single device does not effectively disinfect spaces larger than 4000 cu ft. However, the software allows for synchronous use of multiple AP-4™ devices if larger decontamination room setups are to be considered.

Our study has some limitations. Only one model of N95 FFR was evaluated. There are variations in the construction and materials of different N95 respirators and thus further studies are needed with other N95 models. Our sample size was small, in keeping with the need to preserve N95 FFRs for HCWs. We only evaluated N95 structural integrity and filtration efficiency for up to 5 treatment cycles. Studies are needed to assess the impact of increased number of treatments. Despite these limitations, our study has the advantage of including assessments by a multidisciplinary group which helped evaluate the different factors that would affect the reusability of an N95 FFR.

## CONCLUSION

We found that a PAA room HLDS was effective for the decontamination of N95 respirators with a short cycle time. No adverse effects on filtration efficiency, structural integrity, or strap elasticity were detected after 5 treatment cycles. The PAA room HLDS system provides a rapidly scalable solution for hospitals requiring in-hospital disinfection of N95 respirators.

## Data Availability

All the available data is being made public

## Abbreviations

SARS-CoV-2**=**Severe Acute Respiratory Syndrome Coronavirus; FFR=Filtering facepiece respirator; PAA=Peracetic acid; COVID-19=Coronavirus disease 2019; PPE=Personal Protective Equipment; CDC= Center for Disease Control and Prevention; WHO=World Health Organization; FE=Filtration efficiency; STEL=Short-term exposure limit; TWA=time weighted average; AEGL=Acute exposure guidelines; AGP=Aerosol generating procedure; PUI=Persons under investigation; HCW=Healthcare worker; HLDS=High-level disinfection system; EUA= Emergency Use Authorization.

## Acknowledgement

The authors would like to extend our gratitude to UH Ventures for providing the infrastructure for this study. We would like to thank Altapure Inc™ and ChemDAQ Inc® for their Altapure® AP-4 room disinfection system and Safecide™ E-cell® PAA and hydrogen peroxide remote monitoring system for use in this study respectively. They did not provide input on study design or interpretation of results.

## Contributions

ARJ, SKR and CJD were involved in conceptualizing the study protocols and writing the manuscript; JLC, DFL and SNR performed the inoculation and microbiological reduction analysis; KL helped with the study design, organization of the work, and drawing the images; PM and OA performed the mechanical elasticity testing on the straps; SKM performed the contact angle testing; JW and JAB performed the SEM imaging and structural testing; MB and CKH helped with filtration efficiency testing.

